# Time spent in outdoor light is associated with increased blood pressure, higher hypertension risk, and lower hypotension risk

**DOI:** 10.1101/2024.04.26.24306464

**Authors:** Sicheng Li, Liyong Lu, Wenpan Xian, Jiawei Li, Shuaiming Xu, Jiajin Chen, Yan Wang

**Affiliations:** Xiamen Cardiovascular Hospital of Xiamen University, School of Medicine, Xiamen University, Xiamen, Fujian, China; Center for Health Management and Policy Research, School of Public Health, Cheeloo College of Medicine, Shandong University, Jinan, Shandong, China; NHC Key Lab of Health Economics and Policy Research (Shandong University), Jinan, Shandong, China; West China Hospital of Stomatology, Sichuan University, Chengdu 610041, China; Department of Epidemiology and Health Statistics, West China School of Public Health / West China Fourth Hospital, Sichuan University, Chengdu, Sichuan, China

## Abstract

**Objective:** Light exposure is considered to be associated with reduced blood pressure (BP). However, longitudinal epidemiological studies concerning the light‒BP association with large samples are still limited.

**Methods:** This cohort study enrolled over 300,000 participants from the UK Biobank. Information on time spent in outdoor light during typical summer or winter days was obtained through questionnaires. Cases of hypertension and hypotension were identified using the 10th edition of International Classification of Diseases codes. Cox proportional hazard regression models were employed to estimate the light‒BP associations, restricted cubic splines were utilized to detect potential nonlinear associations, subgroup analyses were conducted to identify effect modifiers, and causal mediation analyses were performed to explore potential mechanisms.

**Results:** Using summer light exposure as an illustration, after a median follow-up of 13.4 years, each additional hour of summer light exposure was associated with an increased risk of hypertension (hazard ratio [HR] 1.011, 95% confidence interval [CI] 1.006‒1.017, *P*-nonlinear=0.803) and a decreased risk of hypotension (0.988, 0.977‒ 0.998, *P*-nonlinear=0.109). The light‒BP association is stronger in females (*P*=0.022), those with short sleep duration (*P*=0.049), and those with high genetic risk of hypertension (*P*<0.001). Potential mechanisms included increasing biological age (proportion mediated, 24.1%, *P*<0.001), neutrophil count (5.4%, *P*<0.001), BMI (32.0%, *P*<0.001), etc.

**Conclusions:** Contrary to previous findings, our study revealed a positive association between light exposure and BP. Potential mechanisms include inflammation, aging, and behavioral lifestyle changes. Further epidemiological and experimental investigations are warranted to validate these novel findings.

## 1. Introduction

Hypertension is the most important modifiable risk factor for global morbidity and mortality, correlating with an increased risk of cardiovascular diseases^1^. According to WHO reports, the number of hypertensive individuals reached 1.3 billion in 2019, doubling from 650 million in 1990, with approximately 33% of individuals aged 30-79 years suffering from hypertension, leading to a significant disease burden^2^. Hypotension, as another type of blood pressure (BP) abnormality, also poses significant health risks that cannot be ignored. For example, orthostatic hypotension has a prevalence ranging from 6% to 35%^3^ and is significantly associated with increased risks of all-cause mortality, coronary heart disease, heart failure, and stroke^4^. Therefore, understanding the risk factors related to BP abnormalities is crucial.

As a common environmental factor, light exposure is considered to be associated with BP. For example, a cross-sectional study involving 5,069 adults nationwide in Chile found that increased geographic latitude was associated with decreased light intensity and increased systolic blood pressure (SBP)^5^. Another cross-sectional study involving 342,457 dialysis patients in the US found that ultraviolet was negatively associated with SBP^6^. The mechanism underlying this association may be that light exposure promotes vitamin D synthesis^7^ and facilitates the release of nitric oxide in the skin into the bloodstream^8^. However, current epidemiological studies have typically employed cross-sectional study designs, which cannot ensure the temporality of exposure and outcome, resulting in relatively low evidence strength, and have not further explored susceptible populations and potential mechanisms of the light‒BP association at the population level.

To address the above gap, this study aims to systematically explore the associations of light exposure with BP, hypertension, and hypotension, identify effect modifiers, and explore potential mechanisms of association based on the baseline survey, repeat survey, and follow-up data from the UK Biobank. Based on existing studies, we hypothesize that light exposure is associated with lower BP, lower risk of hypertension, and higher risk of hypotension.

## 2. Methods

### 2.1 Study population

This study is based on the UK Biobank (www.ukbiobank.ac.uk), a cohort study described in detail previously^9^. In brief, the baseline survey included over 500,000 participants from 37 to 73 from 2006 to 2010, collecting their demographic characteristics, behavioral lifestyle factors, disease history, family history, physical measurements, and biochemical examinations, followed by three repeated surveys including 2012-2013, 2014+, and 2019+. The UK Biobank study has approval from the North West Multi-center Research Ethics Committee, and all participants provided written informed consent for the study^10,11^.

This study included 502,178 participants from UK Biobank. We excluded participants with light exposure time larger than the typical day length in the UK during summer (16h, n=252) and winter (8h, n=5,318)^12^, and participants with extreme light exposure time (larger than the mean plus twice the standard deviation) during summer (n=14,600) and winter (n=24,947)^13^. For hypertension and hypotension outcomes (categorical variables), we separately excluded participants who had the disease before the baseline survey. For the BP outcome (continuous variable), we excluded participants with both hypertension and hypotension before the baseline survey. The design and analysis flowchart of this study is shown in **Figure 1**.

**Figure 1.**
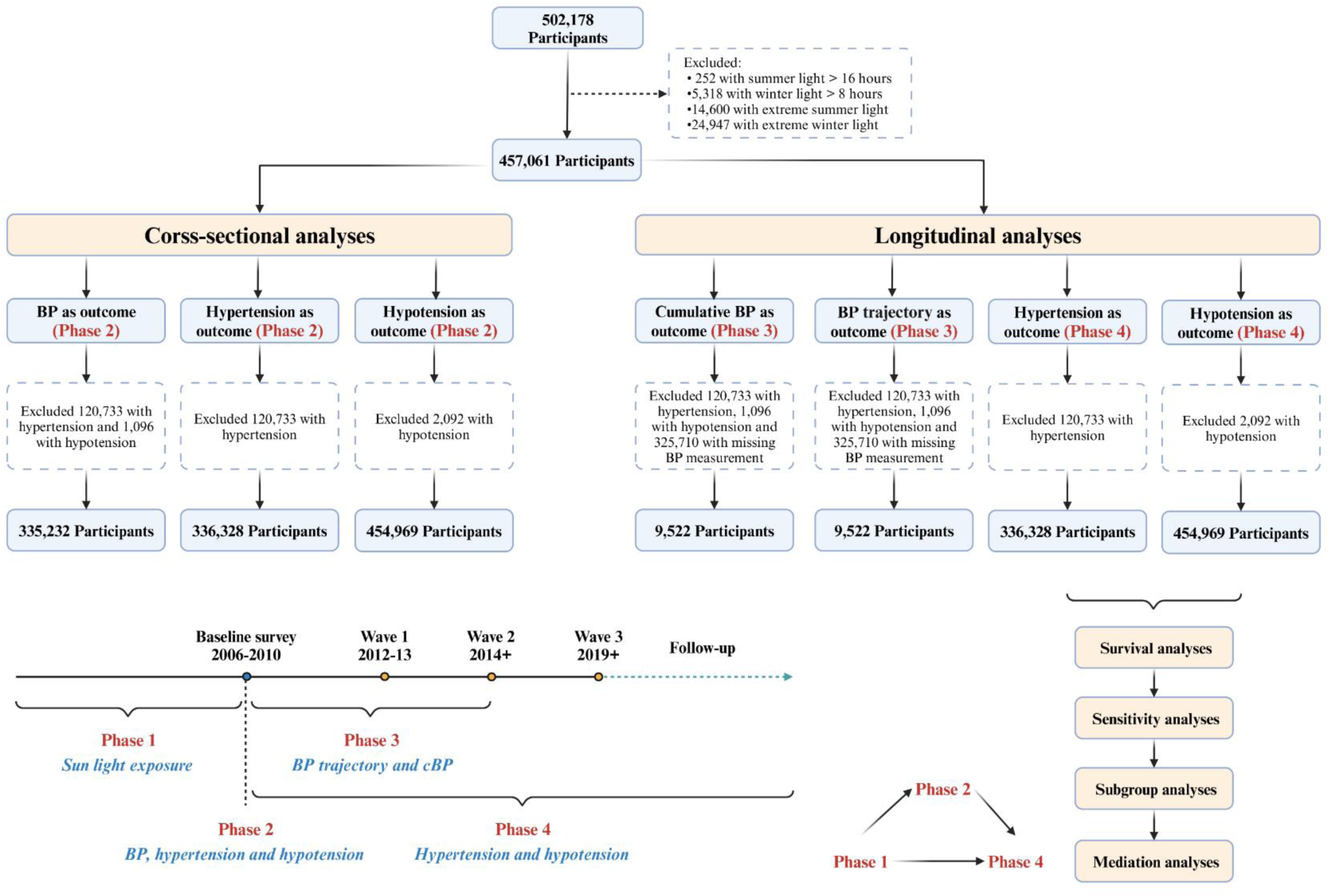
Flow chart of this study. Abbreviations: BP: Blood pressure; cBP: Cumulative blood pressure.

### 2.2 Measurement of light exposure

Participants reported their daily outdoor hours for summer (Field 1050) and winter (Field 1060), including integers or specific values indicating “Do not know”, “Prefer not to answer” or “Less than an hour a day”. The first two specified values are treated as missing data, while the latter is reassigned for half an hour. Additionally, the average daily light exposure was calculated as the mean of summer and winter light, representing the yearly average light exposure.

### 2.3 Measurement of outcome

For BP-associated outcomes in the baseline or repeated survey, the Omron 705 IT electronic BP monitor (df-4079 and df-4080) was used to collect two measures. A manual sphygmometer (df-93 and df-94) was used if the standard automated device could not be employed^14^. We calculated pulse pressure (PP) and mean arterial pressure (MAP) based on systolic blood pressure (SBP) and diastolic blood pressure (DBP), i.e., PP=SBP-DBP, and MAP=(SBP+2×DBP) ÷ 3. The baseline SBP, DBP, PP, and MAP were included as outcomes (**Figure 1, Phase 2**) in the cross-sectional design. Considering the repeated surveys, cumulative BP (cBP, including cSBP, cDBP, cPP, and cMAP) was calculated as the area under the curve of BP (mmHg) versus time (years)^15^. BP trajectory was identified according to the trends of BP utilizing the latent growth model. Analyses involving cBP and BP trajectory (**Figure 1, Phase 3**) included participants with BP data in the baseline and the first two repeat surveys to ensure a relatively sufficient sample size. More details of cBP calculation and BP trajectory identification are shown in **Text S1** and **Figure S1**.

Participants with PB ≥130/80 mmHg and <90/60 mmHg were diagnosed with newly developed hypertension and hypotension (**Figure 1, Phase 2**), respectively, during the baseline survey. The diagnosis of hypertension and hypotension (**Figure 1, Phase 4**) during follow-up utilized the 10th edition of International Classification of Diseases (ICD-10) codes, with the first occurrence recorded in the UK Biobank. Hypertension was identified by ICD-10 codes I10, I11, I12, I13, and I15, while hypotension was identified by code I95. Follow-up concluded at the earliest of the first disease occurrence, death, loss to follow-up, or the end of the follow-up period. The follow-up period was determined as the difference between the baseline survey date and the end of follow-up.

### 2.4 Covariates

We incorporated three sets of variables as covariates: demographic characteristics, behavioral lifestyle, and family history. Demographic characteristics encompassed age (continuous), gender (female, male), education (with or without a college or university degree), employment status (employed, unemployed), skin colour (black, brown, dark olive, fair, light olive, very fair), Townsend Deprivation Index (TDI, continuous), and location (22 assessment centers). Behavioral lifestyle variables included body mass index (BMI, continuous), metabolic equivalent of task (MET, continuous), sleep duration (continuous), healthy diet index (HDI, continuous), smoking status (never, previous, current), drinking status (never, special occasions only, one to three times a month, once or twice a week, three or four times a week, daily or almost daily), sun protection (do not go out in sunshine, never/rarely, sometimes, always, most of the time), and vitamin D supplementation (yes, no). The HDI was derived from participants’ intake of vegetables, fruits, fish, processed meat, and non-processed red meat, with each item scored as 1 for healthy and 0 for unhealthy, yielding an HDI range of 0 to 5, consistent with prior studies^16,17^. Family history was identified according to family history of father, mother, and siblings, including hypertension, diabetes, stroke, and heart disease.

A simplified directed acyclic graph (without considering associations within covariates) was used to depict the relationships among the exposure, covariates, and outcome (**Figure S2**). Demographic characteristics were included as confounders because they were simultaneously associated with light exposure and BP. All these behavioral lifestyle and family history factors were associated with BP. As MET^18^, BMI^19^, and sleep duration^12^ were associated with light exposure, they were included as potential mediators, and the remaining behavioral lifestyle and family history factors were included as effect modifiers.

### 2.5 Statistical analyses

#### 2.5.1 Cross-sectional analyses

In the cross-sectional design, linear regression models were used to estimate the associations (βs) of light exposure with BP, and logistic regression models were used to estimate the associations (odds ratios, ORs) of light exposure with hypertension and hypotension. For each analysis, four models were considered: Model 1, the crude model, where no covariate was included; Model 2, including demographic characteristics; Model 3, further including behavioral lifestyle factors; Model 4, further including family history factors.

#### 2.5.2 Longitudinal analyses (repeated survey)

In the longitudinal design, linear regression models were used to investigate the associations (βs) of light and four cBPs (including cDBP, cSBP, cPP, and cMAP). Compared to the cross-sectional design using BP as the outcome, the advantage of using cBP here is the reduced risk of reverse causality and consideration of BP dynamics, albeit with the disadvantage of a relatively smaller sample size. After the trajectories for the four types of BP (DBP, SBP, PP, and MAP) were successfully clustered, multinomial logistic regression models were used to assess the associations (ORs) between light exposure and the derived BP trajectories. All these models included the same covariates in the above Model 4. More details are shown in **Text S1**.

#### 2.5.3 Longitudinal analyses (follow-up period)

In the longitudinal design, Cox proportional hazard regression models were used to estimate the associations (hazard ratio, HRs) of light exposure with hypertension and hypotension (time-to-event outcome). Restricted cubic spline (RCS) was used to evaluate the nonlinear exposure‒response relationships. Based on the RCS results, continuous variables were transformed into categorical variables to refit the Cox model. Sensitivity analyses conducted to assess the robustness of results included: 1) utilizing multiple imputation techniques (**Text S3**) to address missing data; 2) excluding participants developing hypertension or hypotension within two years after the baseline survey to reduce the risk of causal inversion; 3) replacing the existing hypertension outcomes with primary hypertension (I10); 4) further adjusting fine particulate matter as it may reduce light exposure through physical obstruction^13^ and are associated with BP^20^, potentially acting as a confounder; 5) further adjusting noise and greenspace exposure as light exposure might lead to an increase in these exposures associated with BP^21^, potentially acting as mediators; 6) further adjusting disease such as hearing difficulty and fracture as they might reduce light exposure by spending less time outside and may be associated with BP^13,22,23^, potentially acting as confounders.

To identify susceptible populations, we conducted subgroup analyses by adding interaction terms in the Cox models. Potential effect modifiers included: gender (variables not explicitly annotated here are incorporated in a form consistent with the covariate section), age at baseline (≥60, <60), skin colour, location (England, Scotland, Wales), sun protection, family history of hypertension, sleep duration (>9h, 7-9h, <7h), HDI (divided into three equal parts according to quartiles, i.e., low, medium and high), MET (same as HDI), polygenic score (PRS) for hypertension (divided into five equal parts according to quartiles, with the highest group defined as high risk, the lowest group defined as low risk, and the remaining three groups defined as intermediate risk^9^).

Casual mediation analyses^24^ were conducted to explore potential mechanisms in the light‒BP association (**Text S2**). The mediators predominantly consisted of indicators related to aging, inflammation, lipids, general blood biochemical parameters, and behavioral lifestyle factors. Aging indicators included biological age (BA)^25^, calculated by regressing eight blood indicators on chronological age; phenotypic age, derived by 9 aging-related variables; and telomere length. More details of aging indicators calculation are shown in **Text S3**. Inflammation indicators included C-reactive protein, white blood cell count, neutrophil count, platelet count, lymphocyte count, systemic immune-inflammation index (platelet×neutrophil÷lymphocyte), and bilirubin. Lipids included triglyceride, cholesterol, high-density lipoprotein, low-density lipoprotein, apolipoprotein A, apolipoprotein B, and lipoprotein A. General blood biochemical indicators included calcium, vitamin D, creatinine, urea, glucose, alkaline phosphatase, albumin, hemoglobin A_1c_, Insulin-like growth factors-1, gamma-glutamyl transferase, aspartate aminotransferase, and alanine aminotransferase. Behavioural lifestyle factors included BMI, MET, HDI, and sleep duration.

Participants with missing exposure, outcome, or covariates were further excluded in the main statistical analyses. All analyses were performed using R 4.3.1 software. Two-sided P values ≤0.05 were considered to indicate statistical significance.

## 3. Results

The general characteristics of participants included in the Cox analysis, categorized by hypertension and hypotension, are presented in Table 1. During a median follow-up of 13.4 years (interquartile range, IQR, 12.4‒14.2) and 13.6 years (12.8‒14.3), 61,530 (18.3%) participants and 18,596 (4.1%) participants developed hypertension and hypotension, respectively. Statistical description for light exposure, BP, and cBP is shown in **Table S1**. Taking the population under investigation for the light‒BP association as an example, the distribution of missing data is illustrated in **Figure S3**. A comparison of baseline characteristics of the population before and after participants with missing data were excluded is presented in **Table S2**. Statistically significant changes in baseline characteristics indicate non-random patterns in missing data.

**Table 1.** The general characteristics of the study population.

| Characteristic | Hypertension |  |  | Hypotension |  |  |
| --- | --- | --- | --- | --- | --- | --- |
|  | No<br>N = 274,798, 81.7% | Yes<br>N = 61,530, 18.3% | *P* * | No<br>N = 436,373, 95.9% | Yes<br>N = 18,596, 4.1% | *P* |
| **General CHARACTERISTICS** |  |  |  |  |  |  |
| Age | 54.52 (8.07) † | 58.87 (7.46) | <0.001 | 56.25 (8.07) | 60.97 (6.84) | <0.001 |
| TDI | -1.48 (2.99) | -1.22 (3.12) | <0.001 | -1.37 (3.05) | -0.88 (3.29) | <0.001 |
| Gender (Female) | 167,945 (61.12%) | 32,269 (52.44%) | <0.001 | 249,464 (57.17%) | 9,053 (48.68%) | <0.001 |
| Work (Employed) | 186,227 (68.16%) | 31,417 (51.46%) | <0.001 | 265,933 (61.30%) | 7,126 (38.54%) | <0.001 |
| Education (College or university degree) | 104,125 (38.56%) | 16,877 (28.23%) | <0.001 | 148,753 (34.78%) | 4,616 (25.46%) | <0.001 |
| Location |  |  | <0.001 |  |  | <0.001 |
| England | 242,860 (88.38%) | 55,902 (90.85%) |  | 386,498 (88.57%) | 17,107 (91.99%) |  |
| Scotland | 21,193 (7.71%) | 3,065 (4.98%) |  | 31,953 (7.32%) | 806 (4.33%) |  |
| Wales | 10,745 (3.91%) | 2,563 (4.17%) |  | 17,922 (4.11%) | 683 (3.67%) |  |
| Skin colour |  |  | <0.001 |  |  | <0.001 |
| Black | 1,564 (0.58%) ‡ | 448 (0.74%) |  | 3,160 (0.74%) | 101 (0.56%) |  |
| Brown | 6,857 (2.53%) | 1,938 (3.22%) |  | 12,353 (2.88%) | 486 (2.67%) |  |
| Dark olive | 4,543 (1.68%) | 1,114 (1.85%) |  | 7,485 (1.74%) | 318 (1.75%) |  |
| Fair | 184,037 (67.96%) | 41,271 (68.58%) |  | 292,752 (68.16%) | 12,737 (70.07%) |  |
| Light olive | 52,181 (19.27%) | 10,692 (17.77%) |  | 79,776 (18.57%) | 2,998 (16.49%) |  |
| Very fair | 21,621 (7.98%) | 4,713 (7.83%) |  | 33,977 (7.91%) | 1,537 (8.46%) |  |
| **LIGHT EXPOSURE** |  |  |  |  |  |  |
| Summer light | 3.21 (1.80) | 3.50 (1.88) | <0.001 | 3.32 (1.84) | 3.56 (1.91) | <0.001 |
| Winter light | 1.46 (0.98) | 1.59 (1.05) | <0.001 | 1.51 (1.01) | 1.62 (1.07) | <0.001 |
| Average light | 2.34 (1.24) | 2.55 (1.30) | <0.001 | 2.42 (1.27) | 2.60 (1.31) | <0.001 |
| **LIFESTYLE** |  |  |  |  |  |  |
| BMI | 26.29 (4.25) | 28.21 (4.85) | <0.001 | 27.32 (4.78) | 28.27 (5.44) | <0.001 |
| MET | 2,504.98 (2,418.10) | 2,479.94 (2,492.06) | <0.001 | 2,461.22 (2,420.89) | 2,346.12 (2,487.94) | <0.001 |
| Sleep duration | 7.16 (1.03) | 7.13 (1.17) | <0.001 | 7.15 (1.09) | 7.20 (1.34) | <0.001 |
| HEI | 2.90 (1.28) | 2.83 (1.29) | <0.001 | 2.88 (1.28) | 2.84 (1.28) | <0.001 |
| Smoking status |  |  | <0.001 |  |  | <0.001 |
| Never | 160,881 (58.84%) | 30,641 (50.25%) |  | 242,967 (56.00%) | 8,436 (45.76%) |  |
| Previous | 85,342 (31.21%) | 22,881 (37.52%) |  | 147,877 (34.08%) | 7,492 (40.64%) |  |
| Current | 27,194 (9.95%) | 7,456 (12.23%) |  | 43,045 (9.92%) | 2,507 (13.60%) |  |
| Drinking status |  |  | <0.001 |  |  | <0.001 |
| Never | 19,230 (7.02%) | 5,534 (9.04%) |  | 33,799 (7.77%) | 2,194 (11.86%) |  |
| Special occasions only | 29,077 (10.61%) | 7,750 (12.66%) |  | 49,667 (11.42%) | 2,626 (14.19%) |  |
| One to three times a month | 31,741 (11.58%) | 6,665 (10.89%) |  | 48,902 (11.24%) | 1,966 (10.63%) |  |
| Once or twice a week | 73,059 (26.66%) | 15,118 (24.71%) |  | 112,144 (25.78%) | 4,254 (22.99%) |  |
| Three or four times a week | 66,913 (24.42%) | 13,065 (21.35%) |  | 101,872 (23.42%) | 3,606 (19.49%) |  |
| Daily or almost daily | 53,994 (19.70%) | 13,061 (21.34%) |  | 88,651 (20.38%) | 3,855 (20.84%) |  |
| Sun protection |  |  | <0.001 |  |  | <0.001 |
| Do not go out in sunshine | 1,288 (0.47%) | 498 (0.82%) |  | 2,626 (0.61%) | 211 (1.15%) |  |
| Never/rarely | 22,162 (8.16%) | 6,760 (11.19%) |  | 40,527 (9.40%) | 2,557 (13.97%) |  |
| Sometimes | 87,413 (32.17%) | 20,412 (33.80%) |  | 141,864 (32.92%) | 6,149 (33.59%) |  |
| Always | 57,714 (21.24%) | 12,362 (20.47%) |  | 89,641 (20.80%) | 3,718 (20.31%) |  |
| Most of the time | 103,171 (37.97%) | 20,353 (33.71%) |  | 156,342 (36.27%) | 5,670 (30.98%) |  |
| Vitamin D supplement (Yes) | 11,210 (4.13%) | 2,521 (4.19%) | 0.525 | 17,504 (4.07%) | 816 (4.48%) | 0.006 |
| **FAMILY HISTORY** |  |  |  |  |  |  |
| Family history of diabetes (Yes) | 55,339 (20.49%) | 13,889 (23.18%) | <0.001 | 94,063 (22.02%) | 4,054 (22.59%) | 0.07 |
| Family history of hypertension (Yes) | 120,177 (44.36%) | 27,977 (46.52%) | <0.001 | 210,455 (49.05%) | 8,303 (45.99%) | <0.001 |
| Family history of stroke (Yes) | 64,435 (23.83%) | 17,324 (28.88%) | <0.001 | 114,087 (26.68%) | 5,390 (29.96%) | <0.001 |
| Family history of heart disease (Yes) | 108,471 (40.03%) | 28,345 (47.06%) | <0.001 | 186,948 (43.58%) | 8,932 (49.33%) | <0.001 |

\*: Pearson's Chi-squared test was used for categorical variables, and the Wilcoxon rank sum test was used for continuous variables.
†: Continuous variables are presented as mean (standard deviation), and discrete variables are presented as n (%).
‡: The total number of people here is less than 336,328, as no exclusions for missing data in this table.
Abbreviations: TDI: Townsend deprivation index; BMI: Body mass index; MET: Metabolic equivalent of task; HDI: Health diet index.

### 3.1 Cross-sectional analyses

The findings from cross-sectional analyses are presented in **Table 2**, indicating a significant association of light exposure with elevated BP and increased risk of hypertension, and an insignificant association between light exposure and decreased risk of hypotension. Taking summer light as anexample, exposure to summer light exhibited associations with BP, with effect estimates (βs and 95% CIs) of 0.164 (0.126, 0.201) for SBP, 0.084 (0.063, 0.106) for DBP, 0.079 (0.052, 0.106) for PP, and 0.111 (0.086, 0.136) for MAP. Furthermore, summer light exposure demonstrated associations with hypertension and hypotension, with ORs and 95% CIs of 1.019 (1.014, 1.025) and 0.982 (0.958, 1.005), respectively.

**Table 2.**
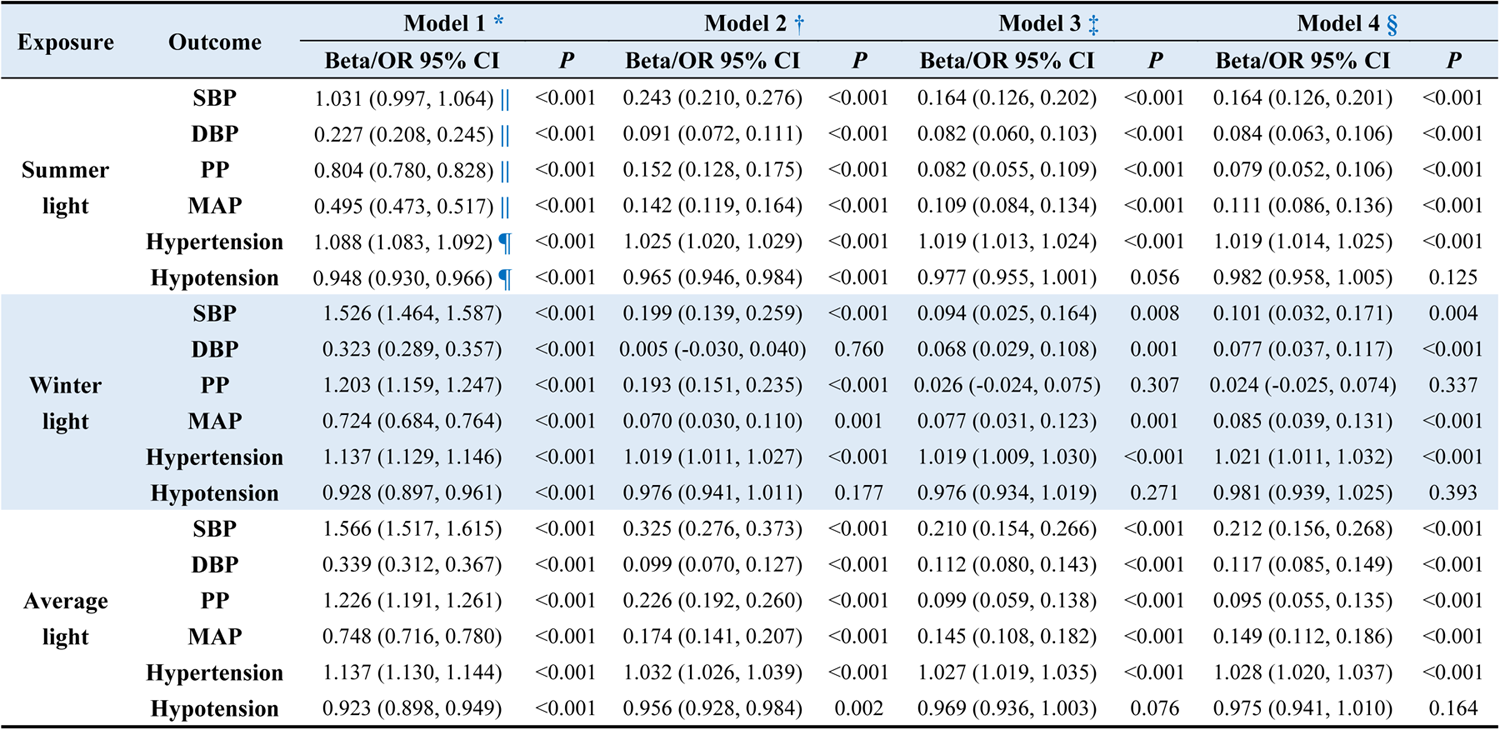
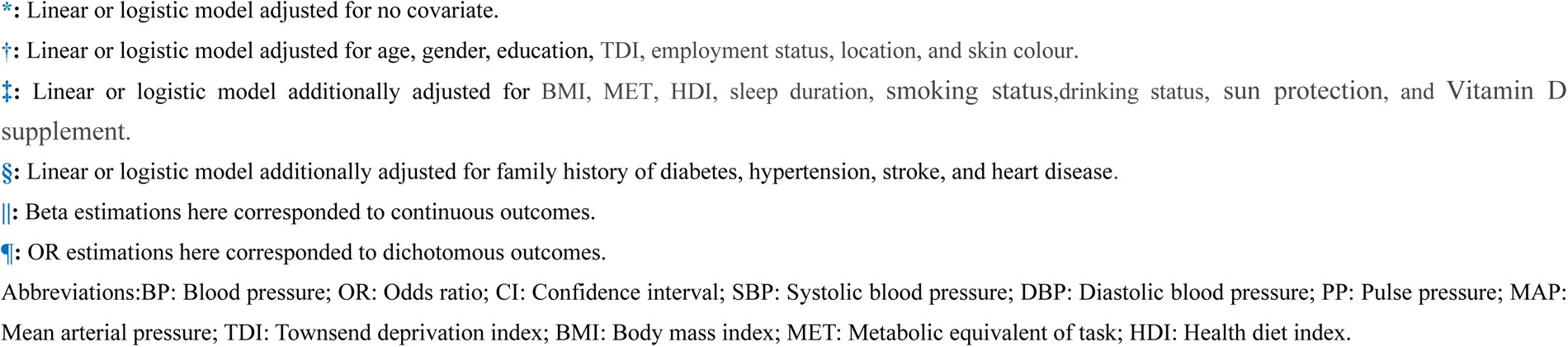
Cross-sectional associations between light exposure and BP.

| Exposure | Outcome | Model 1 * |  | Model 2 † |  | Model 3 ‡ |  | Model 4 § |  |
| --- | --- | --- | --- | --- | --- | --- | --- | --- | --- |
|  |  | Beta/OR 95% CI | P | Beta/OR 95% CI | P | Beta/OR 95% CI | P | Beta/OR 95% CI | P |
| Summer light | SBP | 1.031 (0.997, 1.064) \|\| | <0.001 | 0.243 (0.210, 0.276) | <0.001 | 0.164 (0.126, 0.202) | <0.001 | 0.164 (0.126, 0.201) | <0.001 |
|  | DBP | 0.227 (0.208, 0.245) \|\| | <0.001 | 0.091 (0.072, 0.111) | <0.001 | 0.082 (0.060, 0.103) | <0.001 | 0.084 (0.063, 0.106) | <0.001 |
|  | PP | 0.804 (0.780, 0.828) \|\| | <0.001 | 0.152 (0.128, 0.175) | <0.001 | 0.082 (0.055, 0.109) | <0.001 | 0.079 (0.052, 0.106) | <0.001 |
|  | MAP | 0.495 (0.473, 0.517) \|\| | <0.001 | 0.142 (0.119, 0.164) | <0.001 | 0.109 (0.084, 0.134) | <0.001 | 0.111 (0.086, 0.136) | <0.001 |
|  | Hypertension | 1.088 (1.083, 1.092) ¶ | <0.001 | 1.025 (1.020, 1.029) | <0.001 | 1.019 (1.013, 1.024) | <0.001 | 1.019 (1.014, 1.025) | <0.001 |
|  | Hypotension | 0.948 (0.930, 0.966) ¶ | <0.001 | 0.965 (0.946, 0.984) | <0.001 | 0.977 (0.955, 1.001) | 0.056 | 0.982 (0.958, 1.005) | 0.125 |
| Winter light | SBP | 1.526 (1.464, 1.587) | <0.001 | 0.199 (0.139, 0.259) | <0.001 | 0.094 (0.025, 0.164) | 0.008 | 0.101 (0.032, 0.171) | 0.004 |
|  | DBP | 0.323 (0.289, 0.357) | <0.001 | 0.005 (-0.030, 0.040) | 0.760 | 0.068 (0.029, 0.108) | 0.001 | 0.077 (0.037, 0.117) | <0.001 |
|  | PP | 1.203 (1.159, 1.247) | <0.001 | 0.193 (0.151, 0.235) | <0.001 | 0.026 (-0.024, 0.075) | 0.307 | 0.024 (-0.025, 0.074) | 0.337 |
|  | MAP | 0.724 (0.684, 0.764) | <0.001 | 0.070 (0.030, 0.110) | 0.001 | 0.077 (0.031, 0.123) | 0.001 | 0.085 (0.039, 0.131) | <0.001 |
|  | Hypertension | 1.137 (1.129, 1.146) | <0.001 | 1.019 (1.011, 1.027) | <0.001 | 1.019 (1.009, 1.030) | <0.001 | 1.021 (1.011, 1.032) | <0.001 |
|  | Hypotension | 0.928 (0.897, 0.961) | <0.001 | 0.976 (0.941, 1.011) | 0.177 | 0.976 (0.934, 1.019) | 0.271 | 0.981 (0.939, 1.025) | 0.393 |
| Average light | SBP | 1.566 (1.517, 1.615) | <0.001 | 0.325 (0.276, 0.373) | <0.001 | 0.210 (0.154, 0.266) | <0.001 | 0.212 (0.156, 0.268) | <0.001 |
|  | DBP | 0.339 (0.312, 0.367) | <0.001 | 0.099 (0.070, 0.127) | <0.001 | 0.112 (0.080, 0.143) | <0.001 | 0.117 (0.085, 0.149) | <0.001 |
|  | PP | 1.226 (1.191, 1.261) | <0.001 | 0.226 (0.192, 0.260) | <0.001 | 0.099 (0.059, 0.138) | <0.001 | 0.095 (0.055, 0.135) | <0.001 |
|  | MAP | 0.748 (0.716, 0.780) | <0.001 | 0.174 (0.141, 0.207) | <0.001 | 0.145 (0.108, 0.182) | <0.001 | 0.149 (0.112, 0.186) | <0.001 |
|  | Hypertension | 1.137 (1.130, 1.144) | <0.001 | 1.032 (1.026, 1.039) | <0.001 | 1.027 (1.019, 1.035) | <0.001 | 1.028 (1.020, 1.037) | <0.001 |
|  | Hypotension | 0.923 (0.898, 0.949) | <0.001 | 0.956 (0.928, 0.984) | 0.002 | 0.969 (0.936, 1.003) | 0.076 | 0.975 (0.941, 1.010) | 0.164 |

\*: Linear or logistic model adjusted for no covariate.
†: Linear or logistic model adjusted for age, gender, education, TDI, employment status, location, and skin colour.
‡: Linear or logistic model additionally adjusted for BMI, MET, HDI, sleep duration, smoking status, drinking status, sun protection, and Vitamin D supplement.
§: Linear or logistic model additionally adjusted for family history of diabetes, hypertension, stroke, and heart disease.
||: Beta estimations here corresponded to continuous outcomes.
¶: OR estimations here corresponded to dichotomous outcomes.
Abbreviations: BP: Blood pressure; OR: Odds ratio; CI: Confidence interval; SBP: Systolic blood pressure; DBP: Diastolic blood pressure; PP: Pulse pressure; MAP: Mean arterial pressure; TDI: Townsend deprivation index; BMI: Body mass index; MET: Metabolic equivalent of task; HDI: Health diet index.

### 3.2 Longitudinal analyses (repeated survey)

Light exposure is not significantly associated with cBP. Specifically, exposure to summer light exhibited insignificant associations with cBP, with effect estimates (βs and 95% CIs) of 1.344 (-4.208, 6.897) for cSBP, 0.929 (-2.172, 4.031) for cDBP, 0.415 (-2.290, 3.119) for cPP, and 1.068 (-2.814, 4.949) for cMAP. Based on the combination of the Bayesian information criterion and the interpretability of the trajectory models, the trajectories for four BPs were identified in three categories (**Table S3**). The associations between light exposure and BP trajectories are shown in **Table S4**. Significant associations of summer light and average light with DBP trajectories were found. The increased probability of DBP being classified as an upward-trending trajectory group was significantly associated with summer light (1.094, 1.019‒1.176) and average light (1.141, 1.026‒1.267).

### 3.3 Longitudinal analyses (follow-up period)

The Cox models in this section all satisfied the proportional hazard assumption. The associations of continuous light exposure with hypertension and hypotension are shown in **Table 3**. For example, light exposure was associated with hypertension, with effect estimates (HRs and 95% CIs) of 1.011 (1.006, 1.017) for summer light, 1.008 (0.997, 1.018) for winter light, and 1.015 (1.007, 1.024) for average light. The RCS results are shown in **Figure 2**, where the associations were approximately linear (*P*-nonlinear >0.05) except for the winter light‒hypotension association (*P*-nonlinear <0.05). The associations of categorical light exposure with hypertension and hypotension are shown in **Table 3**, suggesting similar results that much light exposure may elevate the risk of hypertension and reduce the risk of hypotension. Consistent results were observed for sensitivity analyses regarding the association direction, magnitude, and statistical significance (**Tables S5-S10**).

**Figure 2.**
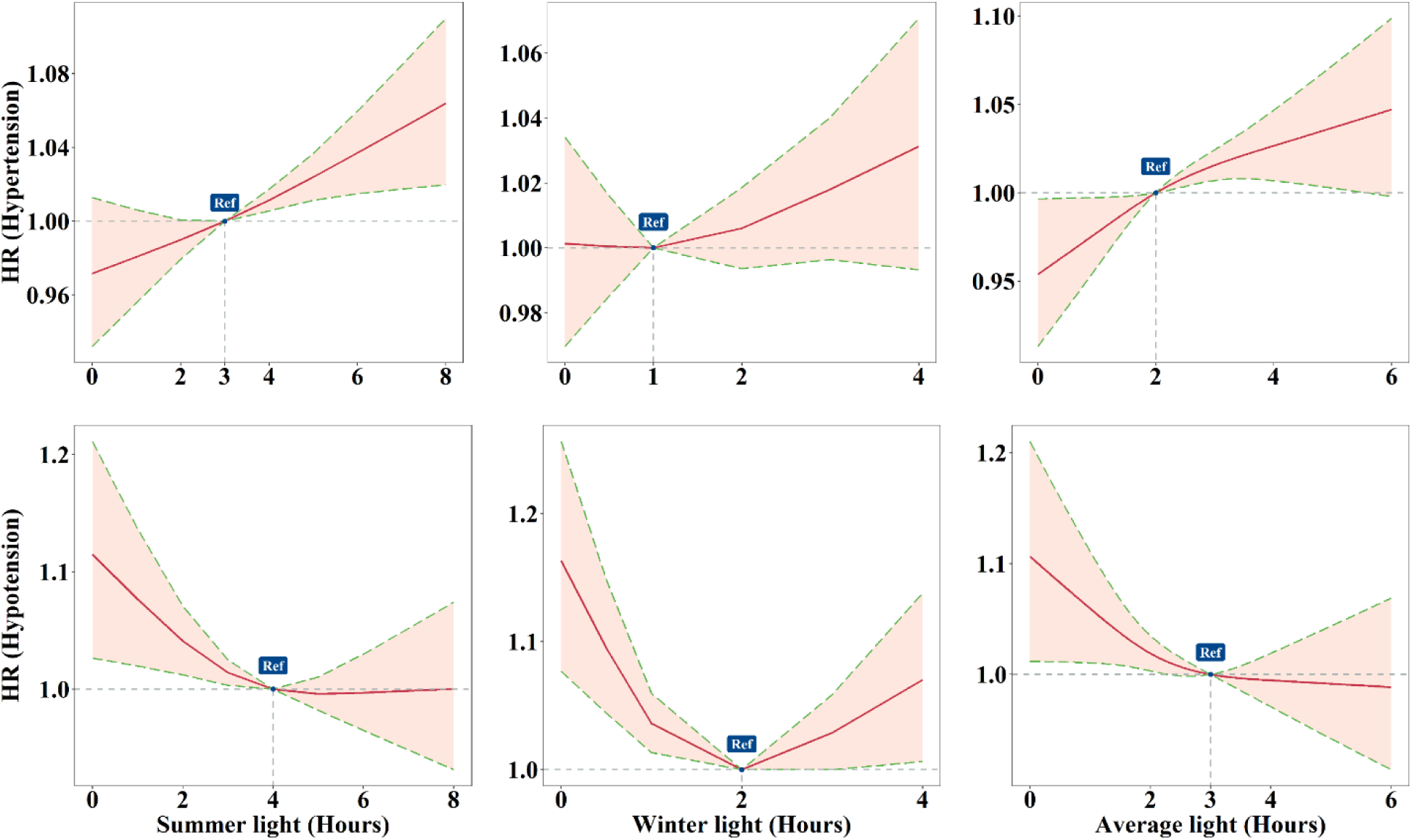
The exposure-response associations of light with hypertension and hypotension. All the models adjusted for age, gender, education, TDI, employment status, location, skin colour, BMI, MET, HDI, sleep duration, smoking status,drinking status, sun protection, Vitamin D supplement, and family history of diabetes, hypertension, stroke, and heart disease. The P-nonlinear from left to right from top to bottom are 0.803, 0.385, 0.471, 0.109, <0.001, and 0.207, respectively. Abbreviations: HR: hazard ratio; TDI: Townsend deprivation index; BMI: Body mass index; MET: Metabolic equivalent of task; HDI: Health diet index.

**Table 3.**
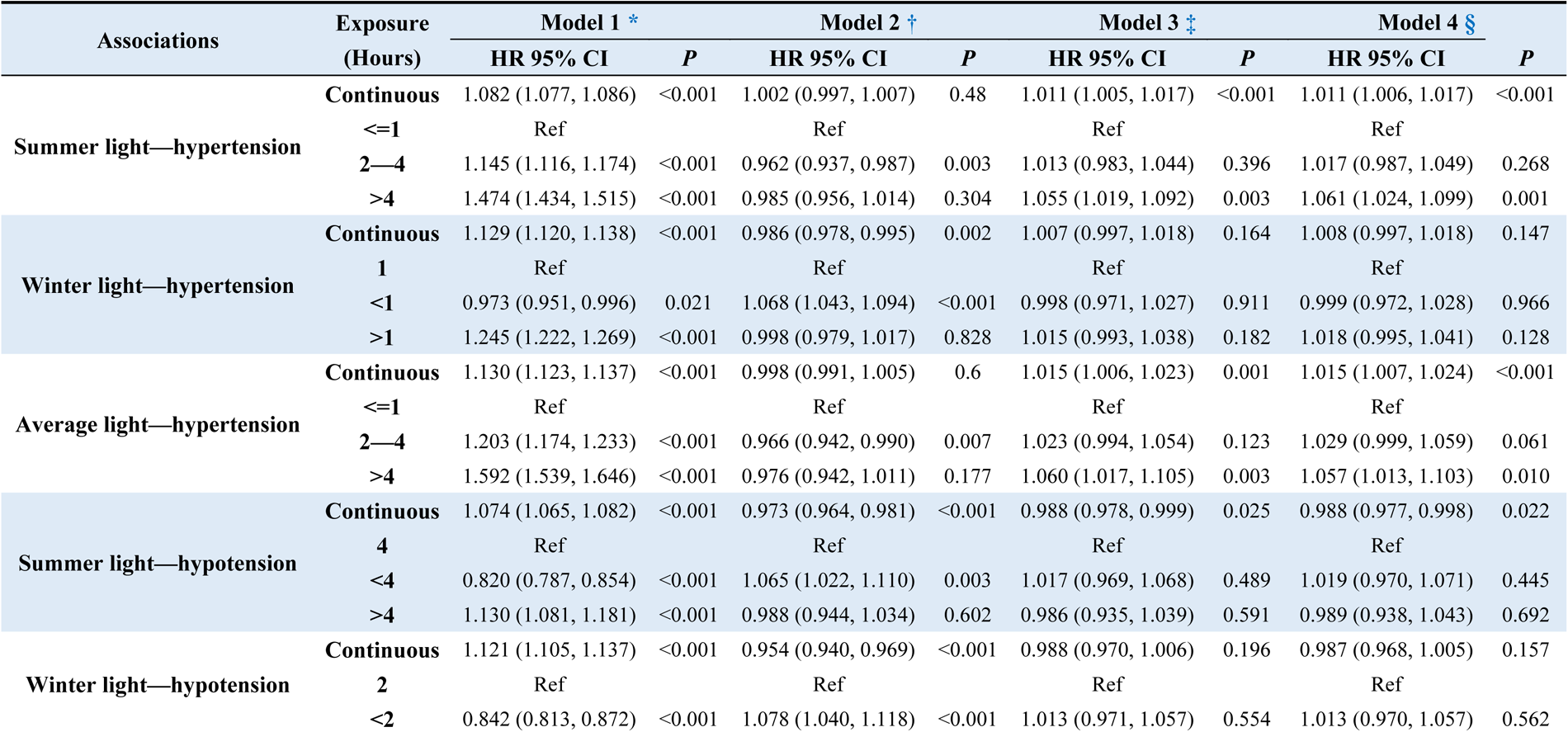
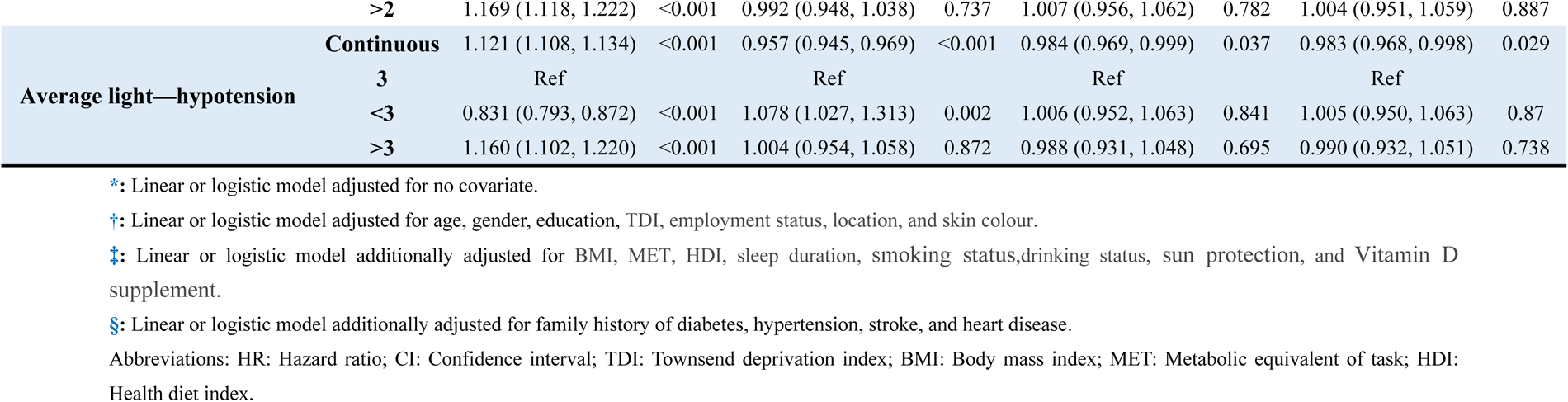
Longitudinal associations of continuous light exposure with hypertension and hypotension.

The subgroup analysis results are presented in **Figure 3**. The main results are summarised as follows. First, Women are sensitive to the BP-raising effects of light exposure compared with men. For example, the summer light‒hypertension association is stronger (*P*=0.022) in females (1.018, 1.010‒1.027) than in males (1.006, 0.998‒1.013); similarly, light exposure provides stronger (*P*=0.012) protection against hypotension in females (0.974, 0.959‒0.989) compared to males (0.998, 0.985‒1.012). Second, longer sleep duration reduces the increased risk of hypertension associated with light exposure and reinforces the benefits of light exposure in lowering the risk of hypotension. For example, the hypertension risk due to average light exposure is higher (*P*=0.049) in the group with short sleep duration (1.020, 1.004‒1.036) than in the group with long sleep duration (0.966, 0.917‒1.017); similarly, hypotension risk decrease due to average light exposure is stronger (*P*<0.001) in the group with long sleep duration (0.859, 0.798‒0.924) than in the group with short sleep duration (1.000, 0.973‒1.029). Third, The genetic risk of hypertension synergistically increases the elevated risk of hypertension due to light exposure, while antagonizing the reduced risk of hypotension due to light exposure. For example, the increased hypertension risk due to summer light is higher (*P*<0.001) in high PRS group (1.085, 1.077‒1.092) than low PRS group (0.929, 0.921‒0.937), while the decreased hypotension risk due to summer light is higher (*P*=0.002) in low PRS group (0.979, 0.965‒0.993) than high PRS group (1.001, 0.987‒1.015).

**Figure 3.**
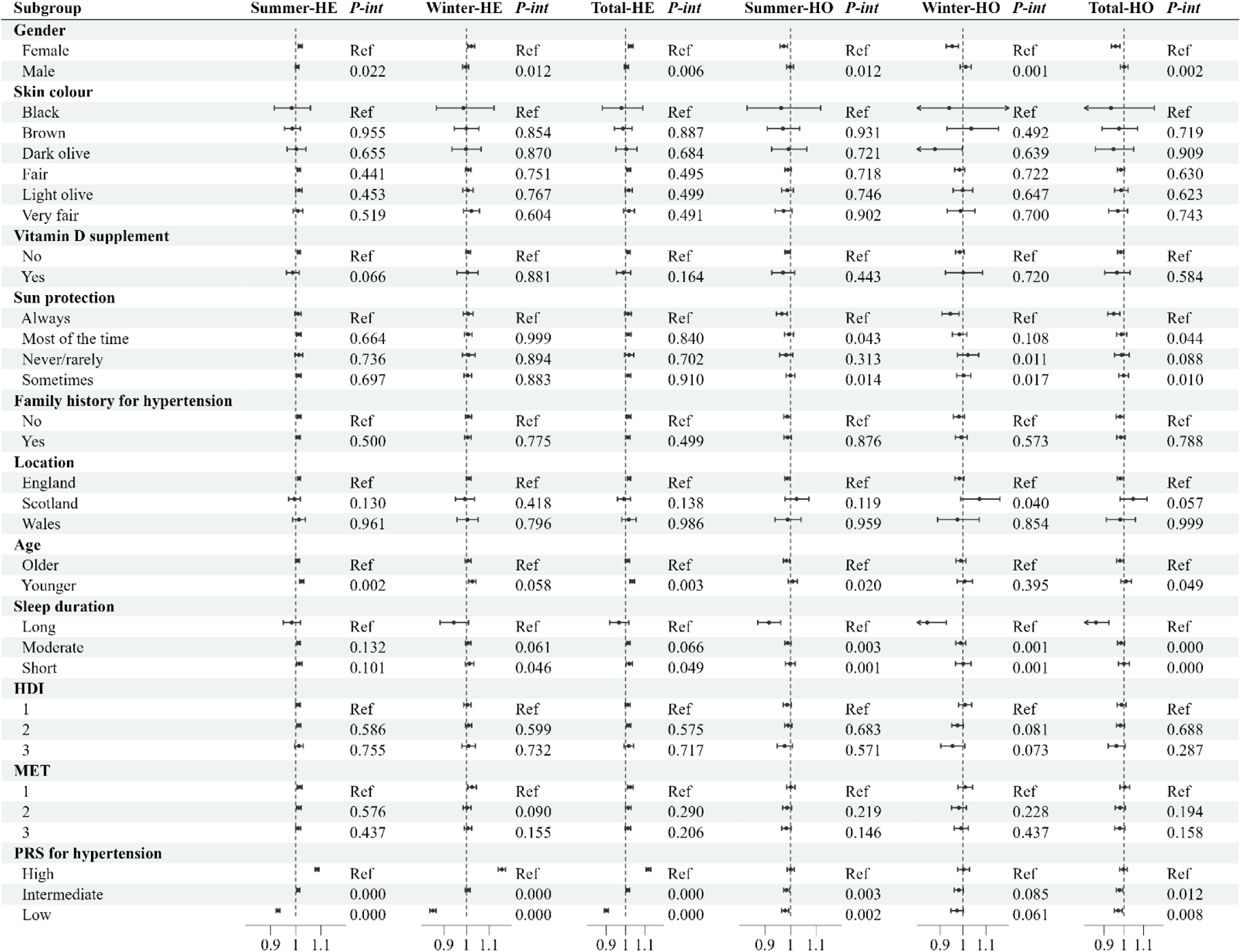
Potential effect modifiers in the associations of light and BP. All the models adjusted for age, gender, education, TDI, employment status, location, skin colour, BMI, MET, HDI, sleep duration, smoking status,drinking status, sun protection, Vitamin D supplement, and family history of diabetes, hypertension, stroke, and heart disease. The effect estimates given in the figure are HRs and 95% CIs. Abbreviations: HE: Hypertension; HO: Hypotension; *P*-int: *P* for interaction; Ref: Reference; HDI: Health diet index; MET: Metabolic equivalent of task; BP: Blood pressure; TDI: Townsend deprivation index; BMI: Body mass index; HR: Hazard ratio; CI: Confidence interval.

The mediation analyses results are demonstrated in **Figure 4**. In brief, significant mediators mainly concentrated on aging, inflammation, and behavioural lifestyle indicators. For example, summer light may increase the risk of hypertension by increasing biological age (proportion mediated, 24.1%, *P*<0.001), neutrophil count (5.4%, *P*<0.001), and BMI (32.0%, *P*<0.001). Summer light associated with reduced risk of hypotension may through increasing MET (39.3%, *P*=0.003).

**Figure 4.**
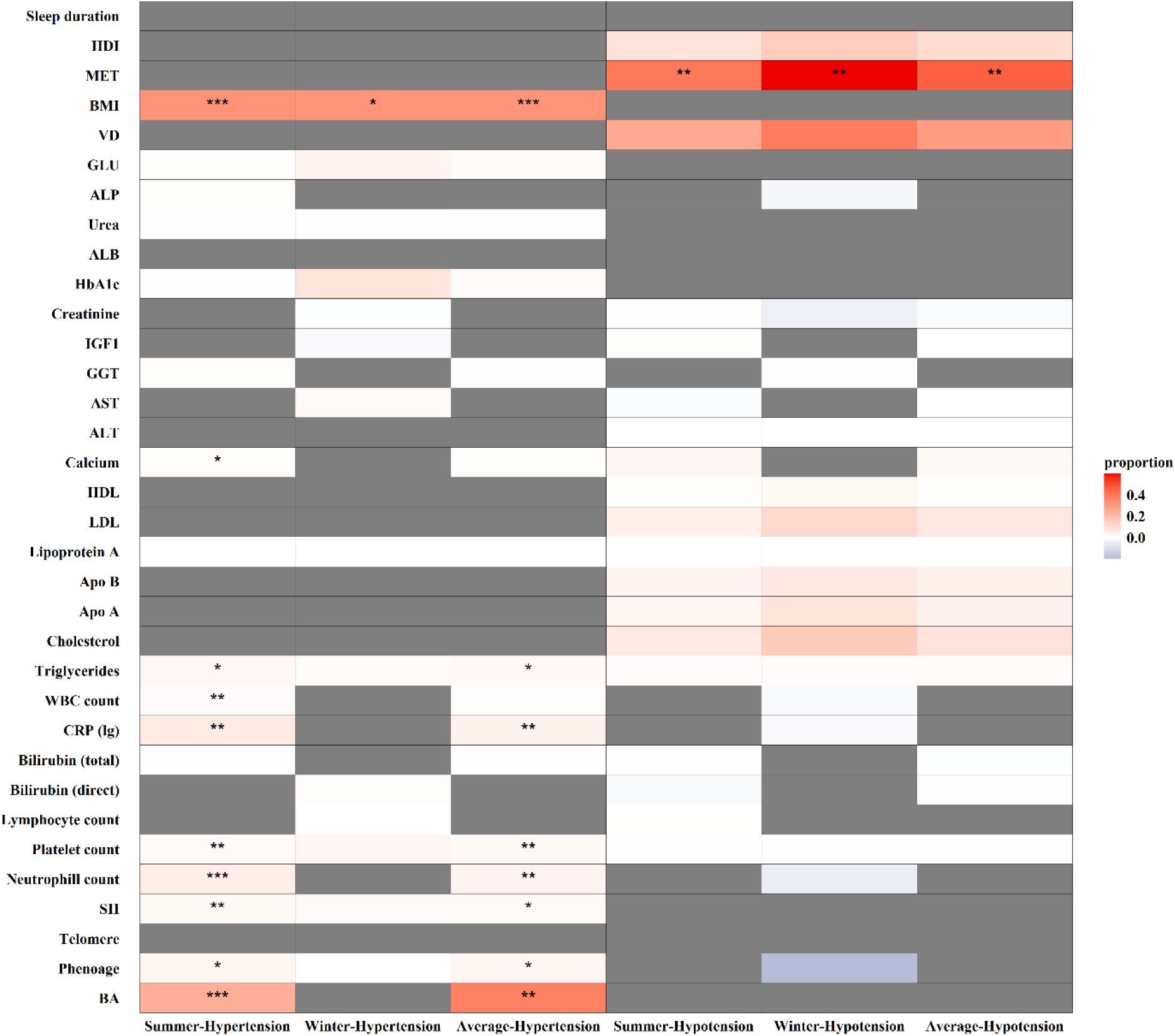
Potential mediators in the associations of light and BP. Both the exposure-mediator and exposure-mediator-outcome models adjusted for age, gender, education, TDI, employment status, location, skin colour, BMI, MET, HDI, sleep duration, smoking status,drinking status, sun protection, Vitamin D supplement, and family history of diabetes, hypertension, stroke, and heart disease. Gray indicates the absence of a mediating effect (total effect and indirect effect are reversed), red indicates a mediating effect by elevating the mediator variable, blue indicates a mediating effect by lowering the mediator variable, and the darker the color, the higher the proportion mediated. Abbreviations: HDI: Health diet index; MET: Metabolic equivalent of task; BMI: Body mass index; VD: Vitamin D; GLU: Glucose; ALP: Alkaline phosphatase; ALB: Albumin; HBA_1c_: Hemoglobin A_1c_; IGF1: Insulin-like growth factors-1; GGT: Gamma-glutamyl transferase; AST: Aspartate aminotransferase; ALT: Alanine aminotransferase; HDL: High-density lipoprotein; LDL: Low-density lipoprotein; APO B: Apolipoprotein B; APO A: Apolipoprotein A; WBC: White blood cell; CRP: C-reactive protein; SII: Systemic immune-inflammation index; BA: biological age; TDI: Townsend deprivation index; √: 0.01< *P* <0.05; ÖÖ: 0.001<*P* <0.01; ÖÖÖ: *P* <0.001

## 4. Discussion

This study represents a pioneering large-scale population-based longitudinal epidemiological investigation into the light‒BP association. Utilizing data from the UK Biobank encompassing over 300,000 individuals, we employed cross-sectional and longitudinal research designs to comprehensively explore the associations of light exposure with continuous BP, hypertension, and hypotension. Additionally, we conducted an in-depth exploration of susceptible populations and potential underlying mechanisms. Our findings indicate that 1) light exposure is associated with elevated BP, increased risk of hypertension, and decreased risk of hypotension; 2) gender, sleep duration, and genetic risk of hypertension may modify the light‒BP association; and 3) mechanisms such as aging, inflammation, and behavioral lifestyle alterations may contribute to the light‒BP association.

### 4.1 Comparison to other studies

Existing epidemiological studies concerning light exposure and BP are limited. A cross-sectional study involving 5,069 adults nationwide in Chile found that increased geographic latitude was associated with decreased light intensity and increased SBP^5^. Similarly, another cross-sectional study involving 342,457 dialysis patients in the US found that ultraviolet A (UVA) and ultraviolet B (UVB) were negatively associated with SBP^6^. Additionally, a cross-sectional study involving 205,888 singleton pregnant women in the US found that higher solar radiation was associated with a lower risk of hypertensive disorders of pregnancy^26^. While these studies suggest a negative relationship between light exposure and BP, contrasting with our findings, differences in study populations may account for this disparity, as our research targeted the general UK population. The strengths of the above studies include the use of geographic coordinates to match participants’ sunlight exposure, ensuring higher accuracy, however without consideration of individual outdoor duration. Our study benefits from a large sample size, self-reported individual outdoor duration, a robust longitudinal design, and comprehensive statistical analyses.

Notably, light exposure is not a singular dimension but comprises complex patterns, including wavelength, intensity, exposure time, and duration^27^. Regarding wavelength, Krause et al. found that UVB, with shorter wavelengths than UVA, is associated with decreased BP, while UVA is not^28^. Concerning intensity, McFadden et al. found a correlation between bedroom brightness and higher BMI in women^29^. In terms of exposure time, Cheung et al. discovered that exposure to blue light in the evening (rather than in the morning) is associated with higher glucose peaks^30^. As for duration, Veleva et al. found that ultraviolet only has short-term effects on BP reduction rather than long-term effects^31^. As a large-scale epidemiological study based on self-reported light exposure, this study accounted for long-term exposure to light encompassing all wavelengths. However, considerations regarding intensity and exposure time remain insufficient in this study.

### 4.2 Potential mechanisms

The potential mechanisms underlying the light‒BP association remain unclear. Existing research supporting the notion that light reduces BP commonly posits that light increases vitamin D synthesis, thereby conferring cardiovascular benefits^7^. However, the relationship between vitamin D and BP remains highly debated^32,33^. A randomized clinical trial involving 119 individuals found that exposure to UVB radiation, which enhances vitamin D synthesis, did not reduce BP^34^. Apart from vitamin D, other studies suggest that light exposure may facilitate the release of nitric oxide from the skin into the bloodstream, thus lowering BP^8,35^.

Mediation analyses suggest that inflammation, aging, and BMI may be potential mediators in light-BP association. Ultraviolet may induce premature skin aging through pathways such as DNA damage, lipid peroxidation, mitochondrial damage, and activation of inflammatory signaling pathways^36^. Furthermore, studies have found that facial aging can predict the risk of cardiovascular disease^37^, possibly due to skin inflammation triggering systemic inflammation and its association with diseases^38^. Since inflammation is a component of aging^39^, facial aging may be related to systemic aging^40^. Regarding lipid metabolism, Klinedinst et al. found that light exposure is associated with decreased subcutaneous fat and increased visceral fat^19^. Pattinson et al. found an association between light exposure and childhood obesity^41^. Similarly, cell experiments have found that ultraviolet-induced adipokines may be associated with decreased subcutaneous fat, leading to disrupted adipose homeostasis and obesity^42^. Animal experiments have also shown that UVB exposure increases food intake and weight gain^43^. The above studies support the role of inflammatory, aging, and BMI in the light-BP association.

### 4.3 Clinical implication

Although the impact of light on BP is relatively modest, the potential risks of hypertension stemming from light exposure cannot be disregarded, given the widespread exposure of the populace to light. The prevalence of individuals exposed to summer light for more than one hour stood at 88.76%, correlating with an HR of 1.041 for hypertension. Consequently, the population attributable fraction amounted to 3.52%, suggesting that a reduction in light exposure to one hour or less among the whole population could alleviate the burden of hypertension by 3.52%. In line with data from the WHO, the global hypertensive population numbered 1.3 billion in 2019. Therefore, curtailing light exposure within one hour could potentially avert approximately 45.76 million cases of hypertension. For susceptible populations, such as females and those with a heightened genetic predisposition to hypertension, as well as vulnerable populations such as outdoor workers exposed to elevated light levels, mitigating light exposure, employing adequate sun protection, supplementing vitamin D, and embracing healthy lifestyles (adequate sleep and low BMI) may prove beneficial.

### 4.4 Limitations and strengths

This study has several limitations. First, this study included self-reported light duration as exposure, which might involve recall bias and change over time, potentially leading to inaccurate effects estimations. In addition, we need more information on light intensity and exposure time to portray light patterns accurately. Second, since self-reported light exposure does not precisely measure individual exposure to light, it may introduce environmental factors such as temperature, humidity, noise, etc., which could affect BP. We controlled some of these factors in our sensitivity analyses and roughly controlled for unmeasured environmental factors by including the UKB assessment centers as a covariate. Third, considering the variable availability and sample size, the covariates included in this study might be insufficient. For example, outdoor physical activity is an unavailable confounder simultaneously associated with light exposure and BP. However, MET was adjusted as an alternative, and we conducted extensive sensitivity analyses regarding covariates and reached consistent conclusions. Fourth, even though this study excluded participants with hypertension and hypotension before the baseline survey, the risk of reverse causality still exists. Consequently, we conducted sensitivity analyses by excluding participants who developed corresponding diseases within two years after the baseline survey and arrived at consistent conclusions, alleviating our concerns. Fifth, the analyses involving cBP and BP trajectory included a relatively small sample size and a limited number of repeated measurements, which may lead to insufficient statistical power. Particularly in trajectory analysis, where samples classified into increasing or decreasing BP groups consist of only a few hundred individuals, the issue of insufficient statistical power may be exacerbated. Finally, missing data is prevalent in this study, which may affect the research findings. However, consistent conclusions were drawn through multiple imputation techniques for missing data. Due to the presence of missing data, the study population is not representative of the UKB population, and considering that the UKB itself may not be representative of the entire population of the UK, caution is needed when extrapolating the findings of this study to the whole UK population.

This study has several strengths. First, we mainly employed a longitudinal study design, decreasing the risk of reverse causality and providing high evidence strength. Moreover, all analyses except for repeated measurements included over 300,000 participants, ensuring sufficient statistical power. Second, comprehensive analyses were conducted on BP, hypertension, hypotension, cBP, and BP trajectories. Consistent results were obtained across these analyses, enhancing the conclusions’ credibility. Finally, numerous sensitivity analyses were conducted to ensure the robustness of the results, susceptible subgroups were identified through subgroup analyses, and potential mechanisms were explored through mediation analyses.

## 5. Conclusion

In conclusion, this study found that light exposure was associated with increased BP, higher risk of hypertension, and lower risk of hypotension. The light‒BP association is stronger among females and those with insufficient sleep or a higher genetic risk of hypertension. Potential mechanisms included inflammation, aging, and behavioral lifestyle changes. Subsequent epidemiological studies considering complex light patterns and in-depth experimental studies are needed to confirm the novel findings of this study.

## Data availability statement

The UK Biobank data can be accessed through approved application (www.ukbiobank.ac.uk)

## Acknowledgments

This study has been conducted using the UK Biobank Resource under Application Number 134551. We express gratitude to all individuals contributed to the acquisition and dissemination of the UK Biobank data, as well as the participants involved in UK Biobank.

